# Generation of Monoclonal Antibodies and Development of Sandwich ELISA for 9-valent HPV Vaccines

**DOI:** 10.1101/2025.07.23.25332104

**Authors:** Shishuai Yan, Baoyang Ruan, Guangjie Tong, Wenwei Wang, Xuanxuan Zhang, Yufeng Cao, Yanrui Yang, Le Sun, Li Shi, Jueren Lou, Maohua Li

**Affiliations:** AbMax Biotechnology Co., Ltd.; Yidao Biotechnology (Suzhou) Co., Ltd.; Shanghai Wellvax Biotechnology Co., Ltd.

**Keywords:** HPV, Neutralizing mA, Sandwich ELISA

## Abstract

Cervical cancer is one of the most common cancers affecting women globally, and nearly all cases of cervical cancer were caused by human papillomavirus (HPV) infection. HPV vaccination has proven to be effective strategy in preventing HPV infection. In this study, for the nine-valent HPV vaccines (HPV 6, 11, 16, 18, 31, 33, 45, 52, and 58), we have generated subtype-specific mouse monoclonal antibodies for all nine HPV subtypes by immunizing the mice with using 9 different virus-like particle (VLP) respectively. Neutralizing monoclonal antibodies specific against each HPV subtypes were identified and used to develop the Sandwich ELISA kits. They can specifically measure the concentration of each subtype VLP in the mixture of 9-valent HPV vaccine for drug substance (DS) or drug product (DP) samples. Most importantly, the ELISA kits can detect the temperature-induced loss of conformational neutralizing antigen epitope(s) in the vaccines, thereby serving as a tool could be used to monitor the stability of the 9-valent HPV vaccines in vitro. In addition, these 9 HPV subtype VLP Sandwich ELISA kits are compatible with HPV VLP vaccines produced by six different vaccines manufacturers, which use three different expression systems with or without the disassembling/reassembling process.

**Highlights:**

1. Subtype-specific HPV VLP Sandwich ELISA kits for the nine-valent HPV vaccines (HPV 6, 11, 16, 18, 31, 33, 45, 52, and 58) were developed using neutralizing monoclonal antibodies
2. These kits can specifically measure the concentration of each subtype VLP in the mixture of 9-valent HPV vaccine for drug substance (DS) or drug product (DP) samples.
3. They can detect the temperature-induced loss of conformational neutralizing antigen epitope(s) in the vaccines, thereby serving as a tool to monitor the stability of the 9-valent HPV vaccines in vitro.
4. These ELISA kits are compatible with HPV VLP vaccines produced by six different vaccines manufacturers, which use three different expression systems with or without the disassembling/reassembling process.

## 1. Introduction

Cervical cancer (CC) stands as a prominent global cause of female mortality, accounting for approximately 530,000 new cases and 275,000 fatalities annually. (1) The development of CC is influenced by multiple risk factors, including high-risk HPV infection, age, smoking, childbirth, oral contraception use, and dietary habits. (2) Amid these variables, persistent infection with high-risk HPV emerges as a pivotal instigator, making CC a growing global public health concern.(3)

The human papillomavirus (HPV) family, embracing over 150 non-enveloped DNA viruses with an affinity for epithelial cells, constitutes a diverse spectrum of types. Among these, over 30 affect the genital tract, and they can be categorized as high-risk or low-risk based on their correlation with CC and precursor lesions. The low-risk types include 6, 11, 42, 43, and 44, while the high-risk types encompass 16, 18, 31, 33, 34, 35, 39, 45, 51, 52, 56, 58, 59, 66, 68, and 70.(4)

First-generation HPV vaccines, such as the bivalent HPV16/18 L1 VLP (bHPV), were designed to target the oncogenic HPV types 16 and 18, which are responsible for approximately 70% of global CC cases. (5) Subsequent advancements led to the development of second generation vaccines, such as the quadrivalent HPV6/11/16/18 L1 VLP (qHPV) and 9-valent HPV 6/11/16/18/31/33/45/52/58 L1 VLP vaccine, which provide broader protection against additional high-risk HPV types (6).

The landscape of CC is undergoing a seismic shift due to vaccine-driven prevention. Clinical trials evidenced the bivalent HPV16/18 vaccine’s capacity to confer ∼90-100% protection against persistent infections and cervical lesions caused by HPV 16 and 18 (7). The quadrivalent HPV6/11/16/18 vaccine demonstrated 100% efficacy in thwarting HPV 16/18-related high-grade cervical, vulvar, and vaginal lesions, while displaying over 90% effectiveness against HPV6/11-linked genital warts. Likewise, the 9-valent HPV 6/11/16/18/31/33/45/52/58 vaccine showcased 97-100% efficacy in forestalling precancerous lesions and persistent HPV infections. It is estimated that the 9-valent vaccine could prevent nearly 90% of HPV-related anogenital cancers, reducing the global disease burden by approximately 600,000 cases annually (8 and 9).

The efficacy of HPV vaccines bears profound significance for manufacturers and recipients alike. Diverse in vitro protocols, encompassing electron microscopy, SDS-PAGE and Western blot, ELISA, L1 binding assays, cell-mediated immunity assessments, and in vitro models, have been employed to evaluate vaccine quality and potency (10). Among these, the sandwich ELISA offers several advantages in the evaluation of the HPV vaccines, such as high sensitivity and specificity, minimal background noise, quantitative analysis, broad applicability, automation and easy-to-use kits, and long-term stability (11). Merck has developed an in vitro potency assessment ELISA kit for their the 4-valent HPV vaccine. The establishment of this method acknowledges the capability of ELISA to validate the efficacy of HPV vaccines in vitro. The results obtained through this sandwich ELISA are correlated with the vaccine’s in vivo activity, protective effects, and immunogenicity, thus laying the foundation for subsequent evaluation and feasibility studies of assay kits designed for assessing HPV vaccines (12). However, Merck’s kits are not commercialized and there is an unmet need for commercially available quantitative ELISA kits to assess HPV vaccine quality across diverse manufacturing processes.

In this study, we describe the development of sensitive, subtype-specific, and universally applicable sandwich ELISA using extensive characterization of HPV subtype-specific monoclonal antibodies. These optimized ELISAs display required specificity, sensitivity, and broad reactivity for a standardized analytical tool kit for monitoring the quality of HPV vaccines.

## 2. Materials and methods

### 2.1 Reagents and suppliers

The HPV16, HPV18, HPV52 and HPV58 virus-like particle (VLP) samples were provided by company 1. And the HPV6, HPV11, HPV31, HPV33 and HPV45 VLP samples were obtained from company 2. Other HPV VLPs are from Chinese vaccine companies, labeled as company 3, company 4, company 5, and so on. Freund’s complete adjuvant (CFA), Freund’s incomplete adjuvant (IFA), Polyethylene glycol 4000 (PEG4000), Dimethyl sulfoxide (DMSO), 3, 3’, 5, 5’-tetramethylbenzidine (TMB) substrates were purchased from Sigma-Aldrich (MO, USA). Female BALB/c mice (4∼6 weeks old) were obtained from Vital River Co. (Beijing, China). Dulbecco’s Modified Eagle Medium (DMEM) and Fetal bovine serum (FBS) were from HyClone (CA, USA). Goat anti-mouse IgG Fc secondary antibodies were purchased from Jackson ImmunoResearch (MA, USA). Horseradish peroxidase (HRP) conjugation kit was purchased from Pierce (IL, USA).

### 2.2 Generation of mouse monoclonal antibodies

As previously described, female BALB/c mice were first immunized with the VLPs of HPV6, HPV11, HPV16, HPV18, HPV31, HPV33, HPV45, HPV52 and HPV58 in Complete Freund’s Adjuvant respectively and boosted in Incomplete Freund’s Adjuvant. Two to four weeks after the first immunization, tail bleeds were tested for titers by indirect ELISA.

The spleen cells isolated from the mice with high antibodies titers were fused with the SP2/0 mouse myeloma cell line. Following fusion, culture supernatants from individual hybridoma clones were screened using ELISA. For antibodies production, the selected hybridoma clones were seeded in stationary bioreactors in DMEM with 10% low-IgG FBS. The bioreactor fluids were collected every 3 days, and IgG fractions were affinity-purified using protein G agarose columns (Cytiva). The concentrations of purified IgG were determined by measuring absorbance at 280nm (OD_280_).

### 2.3 Indirect ELISA

Each well of 96-well high-binding EIA plates was coated with 1000 ng/mL of samples containing different HPV subtype VLPs overnight at 4□ in phosphate-buffered saline (PBS). After two washes with PBS, the wells were blocked with 5% skim-milk in PBS for 1 hour at room temperature. Subsequently, the wells were incubated with either the sera, the culture supernatants, or purified monoclonal antibodies (mAbs) diluted in 5% skim-milk-PBS for 1 hour at room temperature. After two washes with PBS, the wells were incubated with horseradish peroxidase (HRP)-conjugated goat anti-mouse IgG Fc-specific secondary antibodies in 5% skim-milk-PBS for 1 hour at room temperature. After five washes with PBS containing 0.1% Tween20 (PBST), the TMB solution was added. After 30 minutes, the reaction was stopped by adding Stop solution (0.1M H_2_SO_4_), and the absorbance was measured at 450nm using a microplate reader.

### 2.4 Neutralizing antibodies screening

Diluted 293FT cells in complete DMEM medium containing 10% fetal bovine serum, 1% penicillin-streptomycin, 1% L-glutamine, and 1% G418 were seeded into a 96-well cell culture plate and incubated overnight at 37□. The HPV pseudovirus was serially diluted in complete DMEM medium to the desired concentration (approximately 400 fluorescence spots per well). The hybridoma cell supernatants containing the monoclonal antibodies against HPV was initially diluted 20-fold in complete DMEM medium, followed by 4-fold serial dilutions. 60 μL aliquot of the diluted cell supernatants were mixed with an equal volume of the diluted pseudovirus and incubated at 4□ for 1 hour. Subsequently, 100 μL of the mixture was gently added to the 96-well cell culture plate pre-seeded with 293FT cells and incubated at 37□ for 60-96 hours. After incubation, the culture medium was removed, and fluorescence spot counting was performed using an ELISPOT device. The cut-off value was determined as half the number of the fluorescence spots observed in the positive control. Samples with spot counts less than or equal to the cut-off value were considered positive, while those greater than the cut-off were negative.

### 2.5 MAb Pairing and Development of Sandwich ELISA Kits

The antibodies were conjugated with HRP following the manufacturer’s instructions. The specific activities of the conjugated antibodies were determined by calculating the ratio of OD_430_/OD_280_.

Sandwich ELISA. Each well of 96-well high-binding EIA plates (Corning, USA) was coated with different capture mAbs overnight at 4□ in 0.05M Na_2_CO_3_ with pH9.6. After two washes with PBS, the plates were blocked with blocking reagents, then air-dried and sealed in a plastic bag for storage. For antigen detection, the samples were diluted in 3% bovine serum albumin (BSA) in PBS and incubated in the wells for 1 hour at room temperature. After two washes, wells were probed with the second HRP-conjugated mAb diluted in 3% BSA-PBS for another hour. After five washes with PBST, the TMB solution was added to the wells. After 30 minutes, the reaction was stopped by adding Stop solution (0.1M H_2_SO_4_), and the absorbance was measured at 450nm using a microplate reader

Evaluation of the validation parameters of the Sandwich ELISA.

#### Limit of Detection (LOD)

The limit of detection was examined by analyzing the absorbance values of a blank sample (PBS) in replicates. The LOD was defined as being three times the standard deviation;

#### Linearity

It was determined by using the linear correlation coefficient of the standard curves established with the reference vaccine standard. Each point was in triplicates. The linearity was calculated from the linear regression of the results of the standard curve;

#### Specificity

Specificity was determined by testing against the target HPV VLP, the other 8-valent HPV VLP and the 9-valent HPV DS.

#### Precision

Precision was assessed for both intra-assay (within-plate) and inter-assay (between-plate) variability. Vaccine samples with high, medium, and low antigen content were analyzed against the standard curve to evaluate precision. The coefficient of variation (CV) for intra-assay precision was required to be ≤15% and the coefficient of variation (CV) for inter-assay precision was required to be ≤20%.

## 3. Results

### 3.1 Generation of mAbs against the HPV VLPs

To develop a quantitative detection kit for HPV, we first obtained subtype-specific monoclonal antibodies by immunizing mice with VLPs of various HPV subtypes. The hybridoma culture supernatants were initially screened against the corresponding subtype HPV VLP to identify the monoclonal hybridoma cells. These positive clones were expanded and further screened against all nine different subtype HPV VLP to confirm their specification. As shown in Table 1, hundreds of subtype-specific HPV VLP mouse monoclonal antibodies were developed, including 5 mAbs specific for HPV 6, 13 mAbs for HPV 11, 32 mAbs for HPV 16, 37 mAbs for HPV 18, 10 mAbs for HPV 31, 11 mAbs for HPV 33, 15 mAbs for HPV 45, 5 mAbs for HPV 52 and 17 mAbs for HPV 58.

**Table 1:**
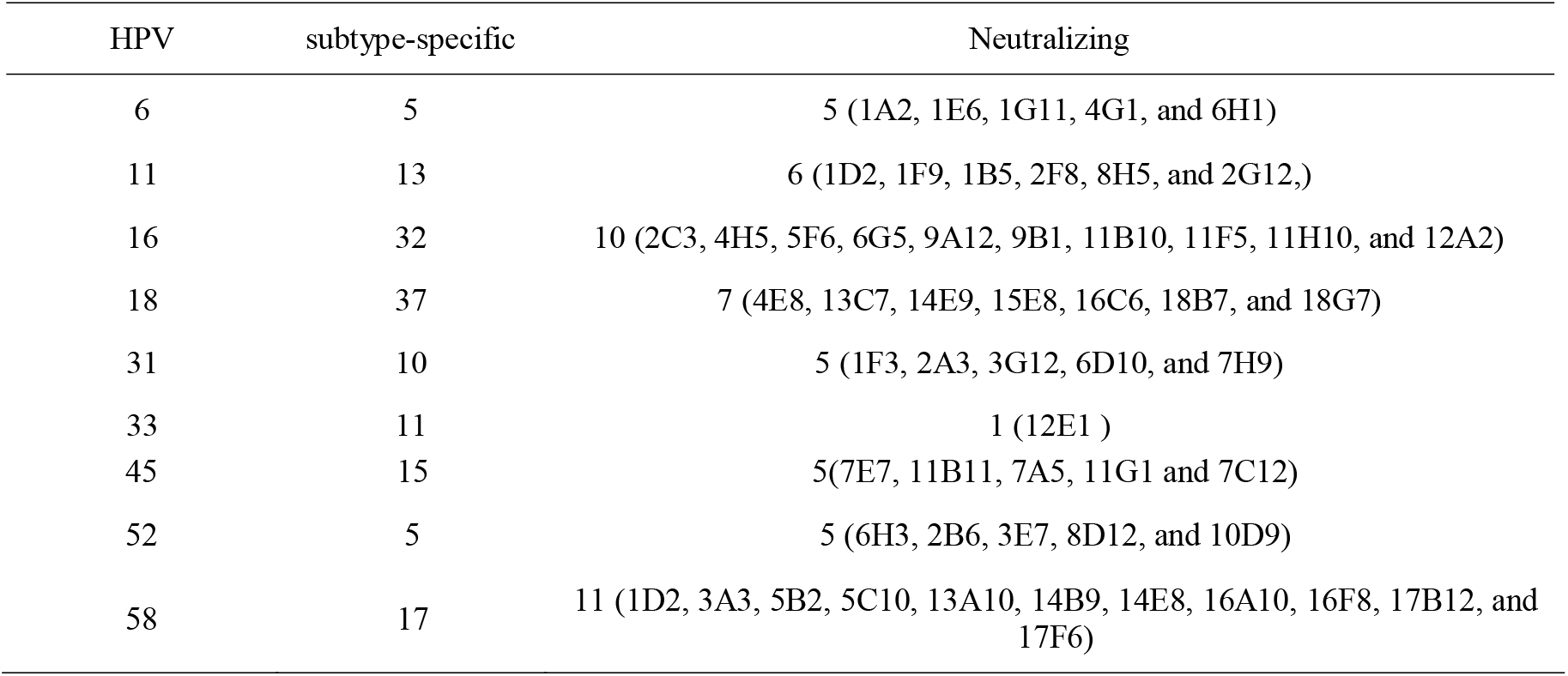
Summary of the mAb obtained with subtype-specific and neutralizing selection.

In hope to develop Sandwich ELISA kits for monitoring both the quantity and the quality of HPV vaccine, all the mAbs were evaluated using an in vitro virus neutralizing assay. Also shown in Table 1, we found more than one neutralizing mAbs for each subtype HPV VLP, except HPV 33 which only has one neutralizing mAb obtained. These neutralizing mAbs are critical for ensuring the functional quality of HPV vaccines, as they reflect the ability of the antibodies to block viral infection in vitro.

### 3.2 mAb pairing

For each HPV subtype, we conducted antibody pairing studies to identify optimal capture and detection antibody combinations for each subtype-specific Sandwich ELISA. Nearly all subtype-specific mAbs were evaluated as either capture and/or detection antibodies. Each well of plates was coated with different mAbs of 5 μg/mL, then paired with HRP-conjugated mAbs of 3 μg/mL to detect HPV VLPs of 2.3, 23 and 230 ng/mL.

Table 2 showed the results of total 20 pairings of 5 neutralizing mAb against HPV45. Among these, the pairing of mAb 7C12 as capturing Ab and mAb 11G1 as detection Antibody gave the best signal-to-noise (S/N) ratio. This combination was selected as the optimal candidate for further development of the HPV45-specific Sandwich ELISA kit.

**Table 2.**
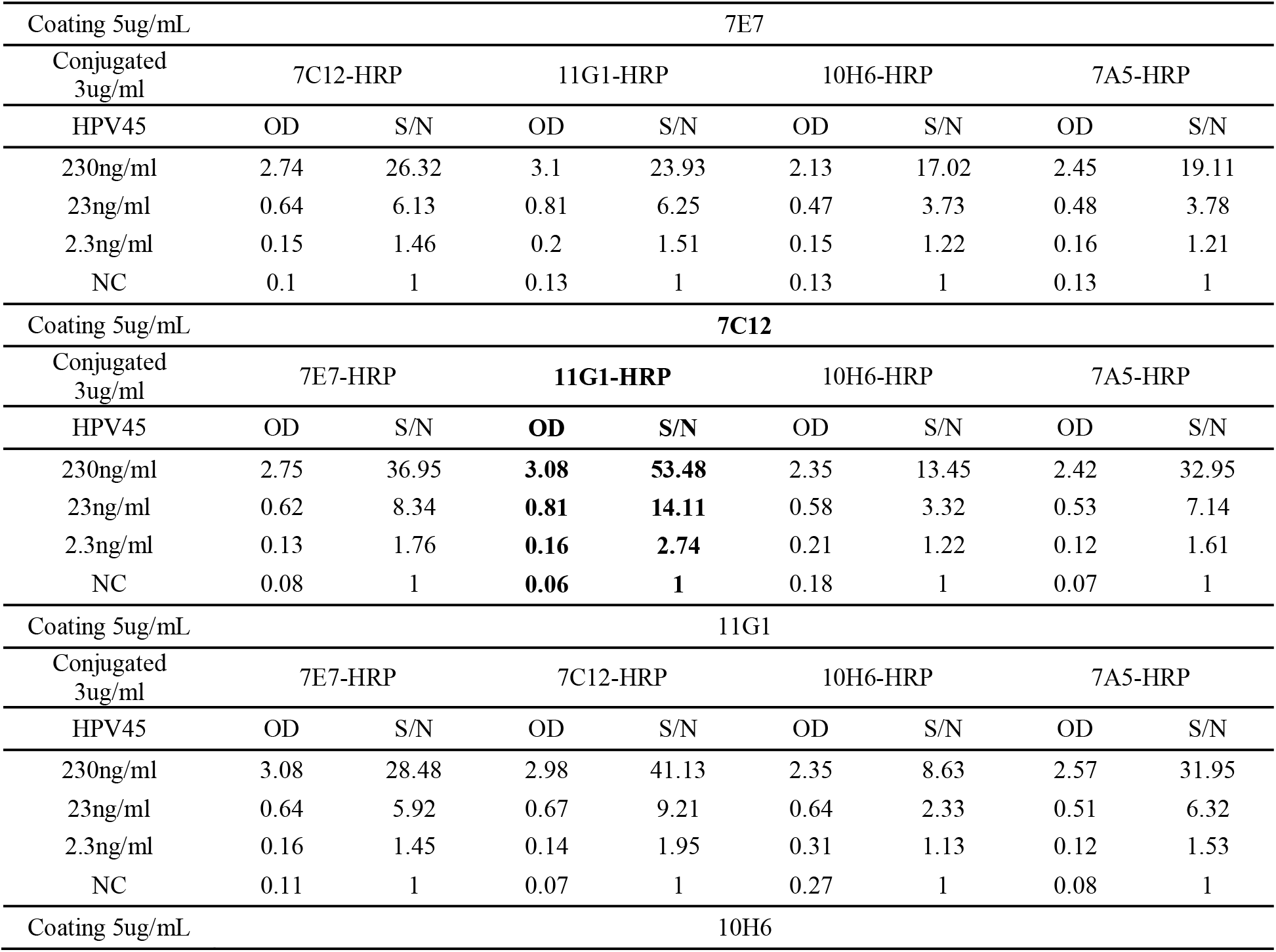
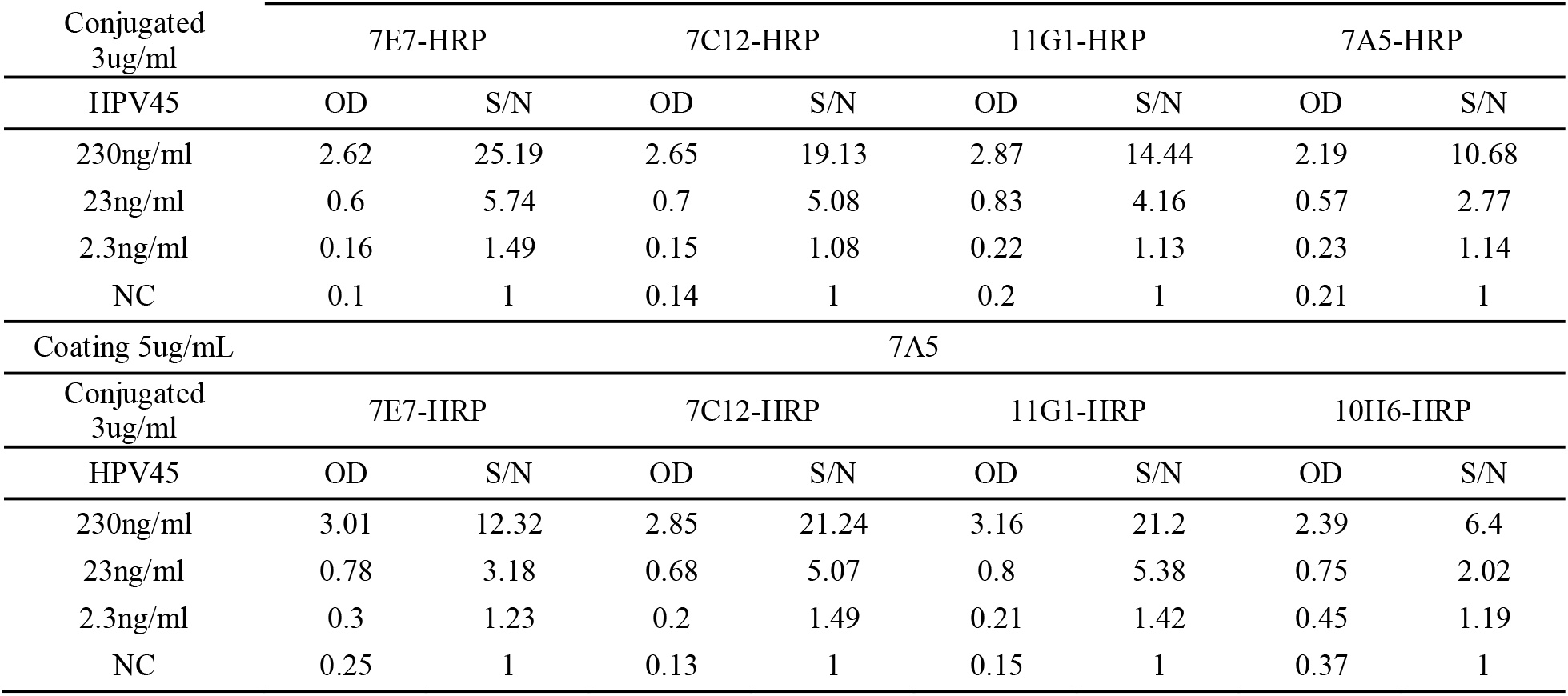
Result of HPV45-specific mAb pairing.

The same systematic mAb pairing tests were carried out for the other 8 HPV subtype VLPs. We identified more than one antibody combination with suitable sensitivity for each of the 9 HPV subtype VLP antigens, as summarized in Table 3.

**Table 3.**
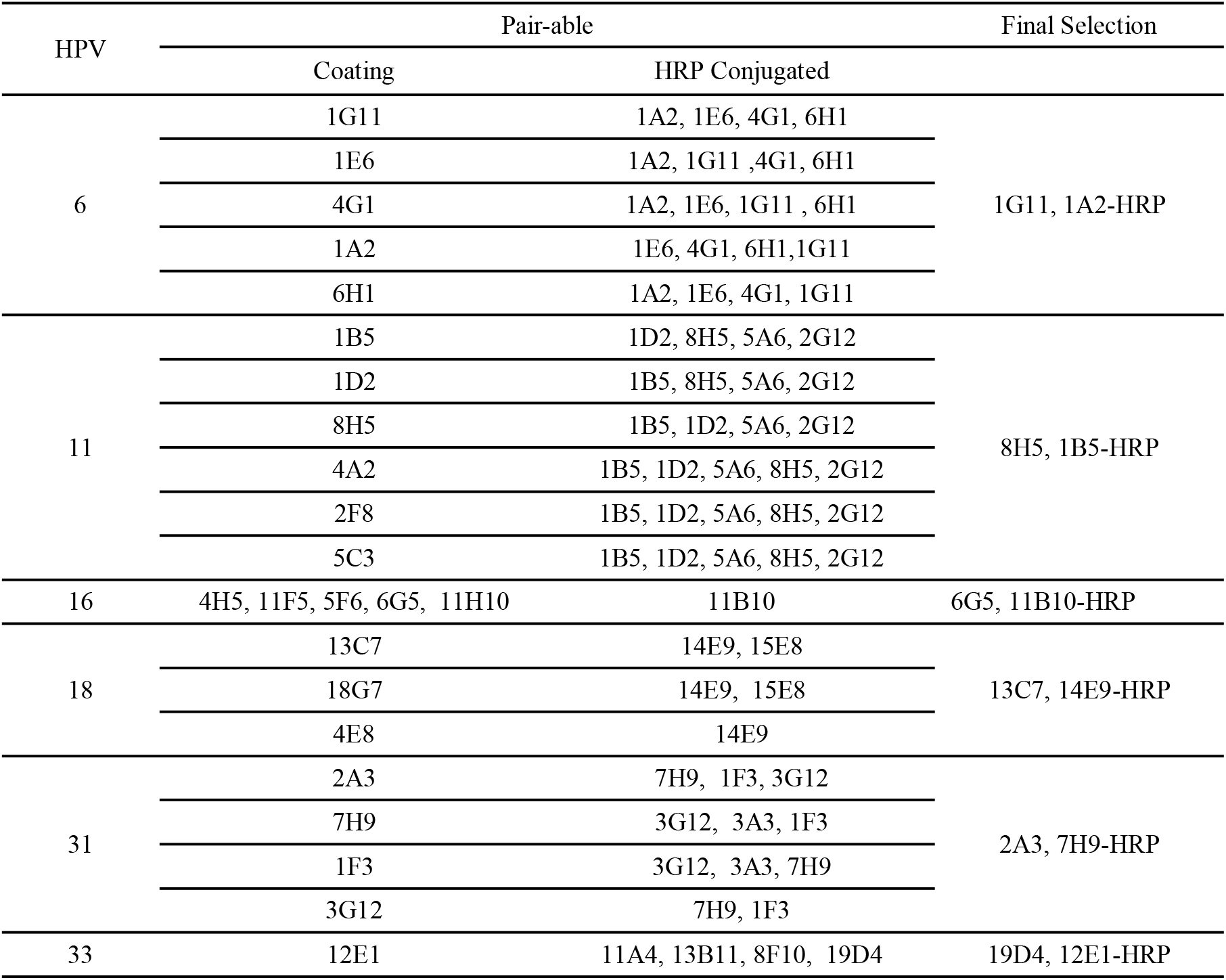
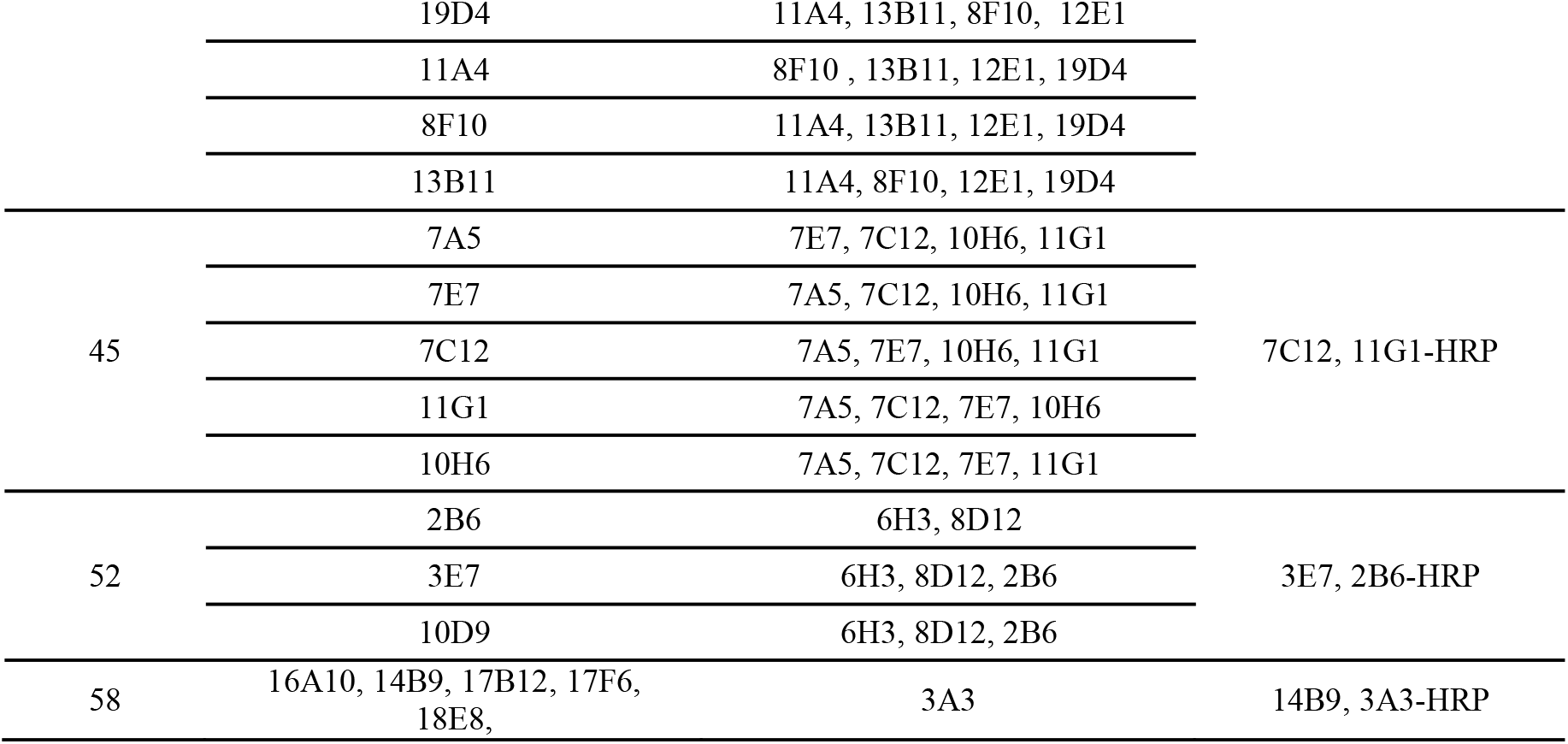
Antibody could be paired detecting targets and the Final selections.

From these tests, the antibody pairings demonstrating the highest analytical sensitivity were selected as candidates for incorporation into the final quantitative ELISA kits, also listed in Table 3. Notably, all kits utilized pairs of neutralizing monoclonal antibodies (mAbs), except for the HPV33-specific Sandwich ELISA kit. For HPV33, the kit employed neutralizing mAb 12E1 as the detection antibody and non-neutralizing mAb 19D4 as the capture antibody. This exception was necessary due to the limited availability of neutralizing mAbs for HPV33, as only one neutralizing mAb (12E1) was identified during screening.

### 3.3 Sandwich ELISA development and manufacturing

#### 3.3.1 Development of Sandwich ELISA

For productions of the 9 assay kits, we tested many manufacture and assay parameters including concentration of capture antibody, concentration of the detection antibody, and so on.

In general, 96-well plates were coated with mAb at five concentrations (0.5, 1, 2, 4 and 8 ug/ml) in 0.05M Na2CO3, pH9.6, overnight at 4□. After washing, the plates were blocked with 3% BSA in PBS, and then air-dried. Twelve different concentrations of vaccine standards were added into each well of the plate, detected with three different concentrations of HRP-conjugated mAb.

As summarized in Fig. 1 and Table 4, All standard curves were fitted using four-parameter logistic model, y=A2+(A1-A2)/(1+(x/x0)^p. And all of the 9 kits exhibited a broad linear range of 3.9-4000 ng/mL VLP antigen with LOD at 3.9 ng/mL or better and R^2^ was more than 0.99. These enables accurate measurement of the individual VLP components present in HPV vaccines, which are formulated in the range of 10-40 ug/0.5mL for each VLP subtype.

**Table 4:** 4-Parameters Standard Curves.

| Subtype | A1 | A2 | x0 | p | R2 | LOD |
| --- | --- | --- | --- | --- | --- | --- |
| 6 | 0.11039 | 4.23680 | 139.13636 | 1.50616 | 0.99470 | 3.9ng/mL |
| 11 | 0.03260 | 3.83499 | 364.11174 | 1.19729 | 0.99940 | 3.9ng/mL |
| 16 | 0.10928 | 4.21738 | 124.38040 | 1.28896 | 0.99811 | 3.9ng/mL |
| 18 | 0.05292 | 4.24212 | 251.46513 | 1.00054 | 0.99842 | 3.9ng/mL |
| 31 | 0.11782 | 4.31597 | 276.77958 | 1.37593 | 0.99726 | 3.9ng/mL |
| 33 | 0.06777 | 4.21677 | 152.98954 | 1.27598 | 0.99876 | 3.9ng/mL |
| 45 | 0.02543 | 4.00641 | 120.55552 | 1.06357 | 0.99909 | 3.9ng/mL |
| 52 | 0.12425 | 4.23063 | 379.81190 | 1.09024 | 0.99949 | 3.9ng/mL |
| 58 | 0.07118 | 4.15971 | 216.74950 | 1.06771 | 0.99943 | 3.9ng/mL |

**Figure 1:**
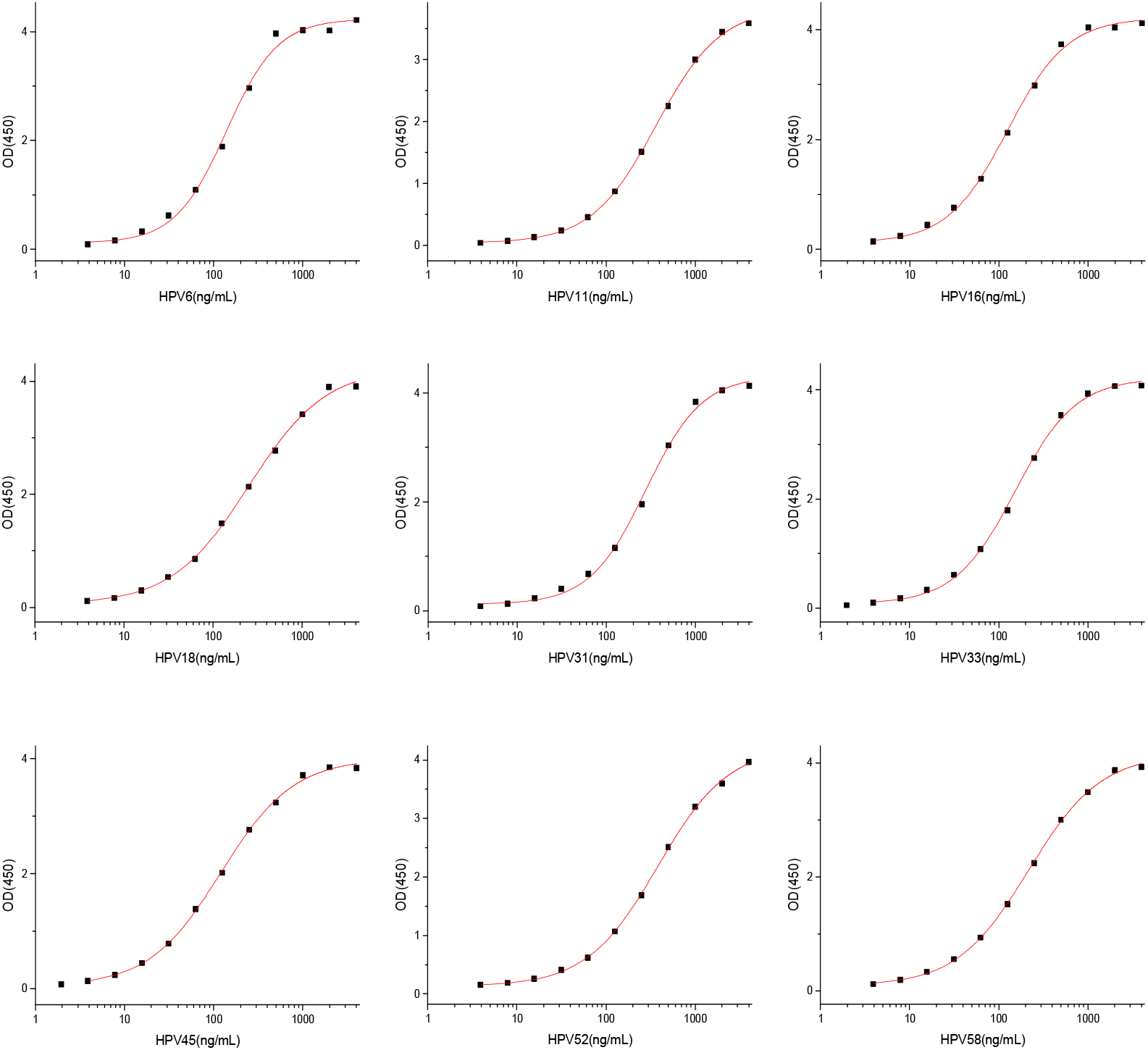
The Standard Curves for 9 different HPV VLP Quantitation Assays

#### 3.3.2 Specificity

Specificity was evaluated by testing each ELISA kit against the target HPV VLP, the other 8-valent HPV VLP and the 9-valent HPV DS.

As summarized in figure 2, using HPV 6 VLP Quantitation Sandwich ELISA kit as an example, the testing samples were 1) HPV 6 VLP only, 2) mixture of 8 different VLPs (HPV 11, 16, 18, 31, 33, 45, 52, and 58), 3) 9-valent HPV DS (HPV 6, 11, 16, 18, 31, 33, 45, 52, and 58). No significant cross-reactivity was observed with the mixture of 8 different VLPs (HPV 11, 16, 18, 31, 33, 45, 52, and 58) (line with dot) over a range from 3.9-500 ng/mL of each HPV VLP. At the same time, the recovery of HPV 6 in the 9-val HPV DS (line with triangle) is above 90%, as evidenced by the overlapping of the two standard curves between mono HPV 6 and 9-val HPV DS. Similar high specificity for the other 8 different HPV subtype VLP Sandwich ELISA kits were also demonstrated. These results confirmed the robust specificity of the ELISA kits for their respective target HPV subtypes, with minimal cross-reactivity and high accuracy in complex mixtures.

**Figure 2:**
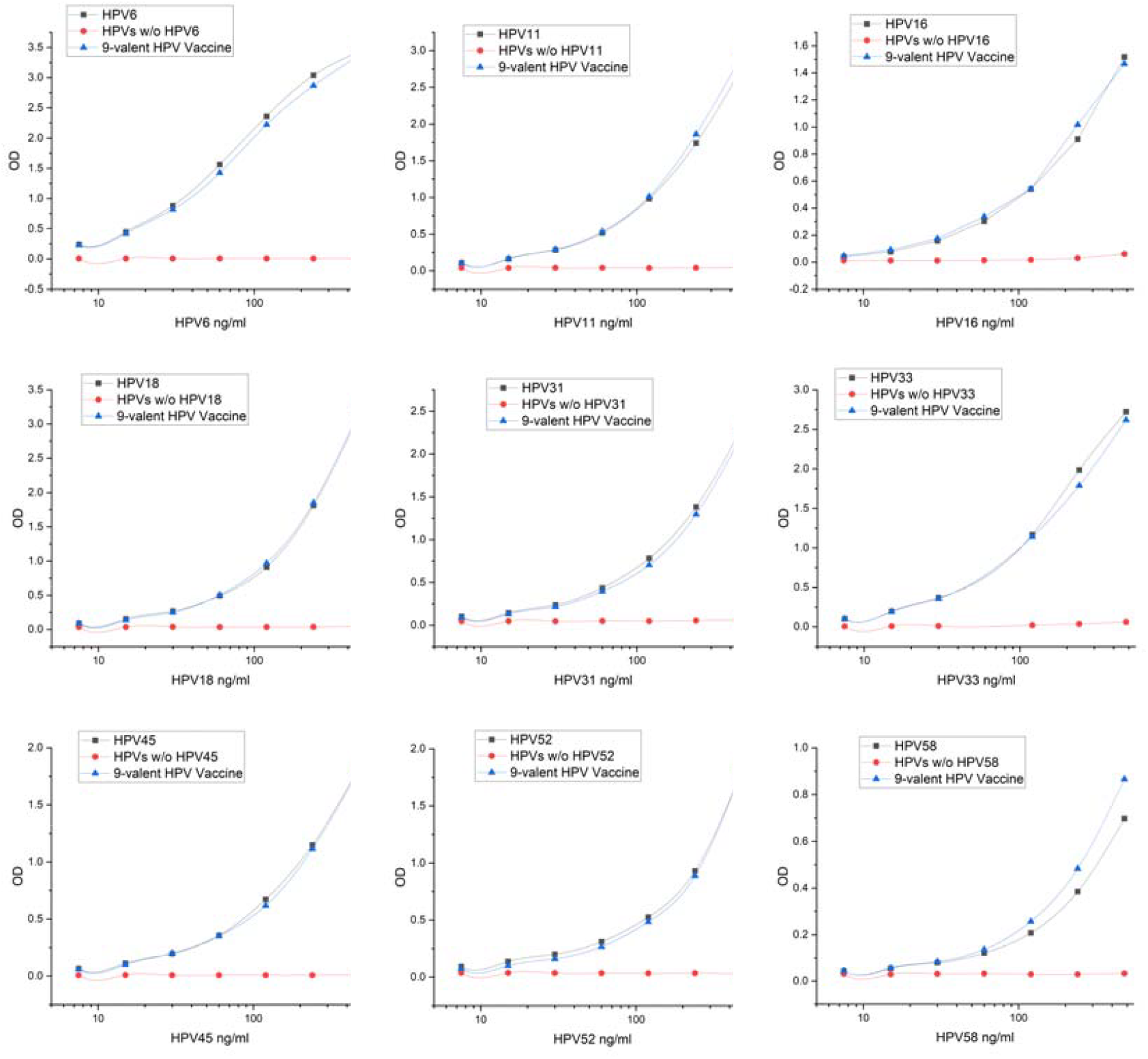
Specificity of 9 different HPV subtype VLPs.

#### 3.3.2 Precision

Precision was assessed for both intra-assay (repeatability) and inter-assay (intermediate precision) variability. To evaluate inter-assay precision, the ELISA was performed by multiple technicians, and the standard deviation (SD) and coefficient of variation (CV) were calculated.

As shown in Table 4, the intra-assay CV ranged from 2.5% to 5.2%, while the inter-assay CV ranged from 2.7% to 5.8%. These results demonstrate the high reproducibility and reliability of the assays across different runs and operators.

**Table 4.** Precision of the Assays.

| Sample | Intra-Assay | Inter-Assay |
| --- | --- | --- |

|  | SD | CV | SD | CV |
| --- | --- | --- | --- | --- |
| sample 1 | 5.2485 | 5.2% | 2.8313 | 2.7% |
| sample 2 | 1.1599 | 5.0% | 0.7919 | 3.4% |
| sample 3 | 0.1623 | 2.5% | 0.3886 | 5.8% |

### 3.4 Heat treatment dramatically reduces the ELISA titers

Since every kits incorporate at least one HPV subtype-specific neutralizing antibodies, these assays were capable of detecting immunologically relevant epitopes on the VLPs that elicit protective antibody responses in vivo.

To evaluate the utilities of the HPV VLP Sandwich ELISA kits for monitoring product stabilities, HPV VLP samples were heat-inactivated at 56□ for 30 minutes, followed by cooling to room temperature. Heat-treated and untreated VLPs were measured by the ELISA kits. As shown in Table 5, heat treatment dramatically reduced the ELISA signals, indicating the loss of conformational neutralizing epitope(s) in the VLPs. This suggested that the Sandwich ELISA kits could be further developed into the in vitro potency assays for human HPV vaccines, providing a reliable method to access vaccine stability and functional integrity.

**Table 5.** Reducing Rate of the recognition of Heat treated VLPs sample with the untreated.

| HPV6 |  |  |  | HPV11 |  |  | HPV16 |  |  |
| --- | --- | --- | --- | --- | --- | --- | --- | --- | --- |
| ng/m<br>L | Un-Treated | Treat-<br>ed | Reducing<br>Rate | Un-Treated | Treat-<br>ed | Reducing<br>Rate | Un-Treated | Treat-<br>ed | Reducing<br>Rate |
| 240 | 2.097 | 0.027 | 98.71% | 1.874 | 0.021 | 98.88% | 1.549 | 0.035 | 97.74% |
| HPV18 |  |  |  | HPV31 |  |  | HPV33 |  |  |
| ng/m<br>L | Un-Treated | Treat-<br>ed | Reducing<br>Rate | Un-Treated | Treat-<br>ed | Reducing<br>Rate | Un-Treated | Treat-<br>ed | Reducing<br>Rate |
| 240 | 3.486 | 0.23 | 93.40% | 2.233 | 0.165 | 92.61% | 1.877 | 0.073 | 96.11% |
| HPV45 |  |  |  | HPV52 |  |  | HPV58 |  |  |
| ng/m<br>L | Un-Treated | Treat-<br>ed | Reducing<br>Rate | Un-Treated | Treat-<br>ed | Reducing<br>Rate | Un-Treated | Treat-<br>ed | Reducing<br>Rate |
| 240 | 1.469 | 0.12 | 91.83% | 2.086 | 0.058 | 97.22% | 0.491 | 0.045 | 90.84% |

Next, we prepared in-activated HPV 16 VLP samples by mixing different ratio of active VLP samples with Heat-inactivated VLP samples, 100% active (square), 20% active (triangle) and totally inactive (circle). One hand, we used the three samples to immunize different groups of mice, in the presence of adjuvant. On day 28 after immunization, the mouse sera were collected and the total antibody titers were measured against the HPV 16 VLP by indirect ELISA. On the other hand, the three modified HPV 16 VLP samples were also measured by the HPV 16 VLP Sandwich ELISA. As shown in Fig. 3-1, the total anti-HPV 16 VLP antibody titers could not identify the loss of neutralizing antigenic epitope(s) by heating, while the HPV16 VLP Sandwich ELISA kit could accurate detect the an 80% loss of ELISA signal in the 20% active sample (Fig. 3-2). Our data demonstrate the HPV 16 VLP Sandwich ELISA kit was a more sensitive, more accurate, faster and simple assay to assess the stability of the product.

**Figure 3:**
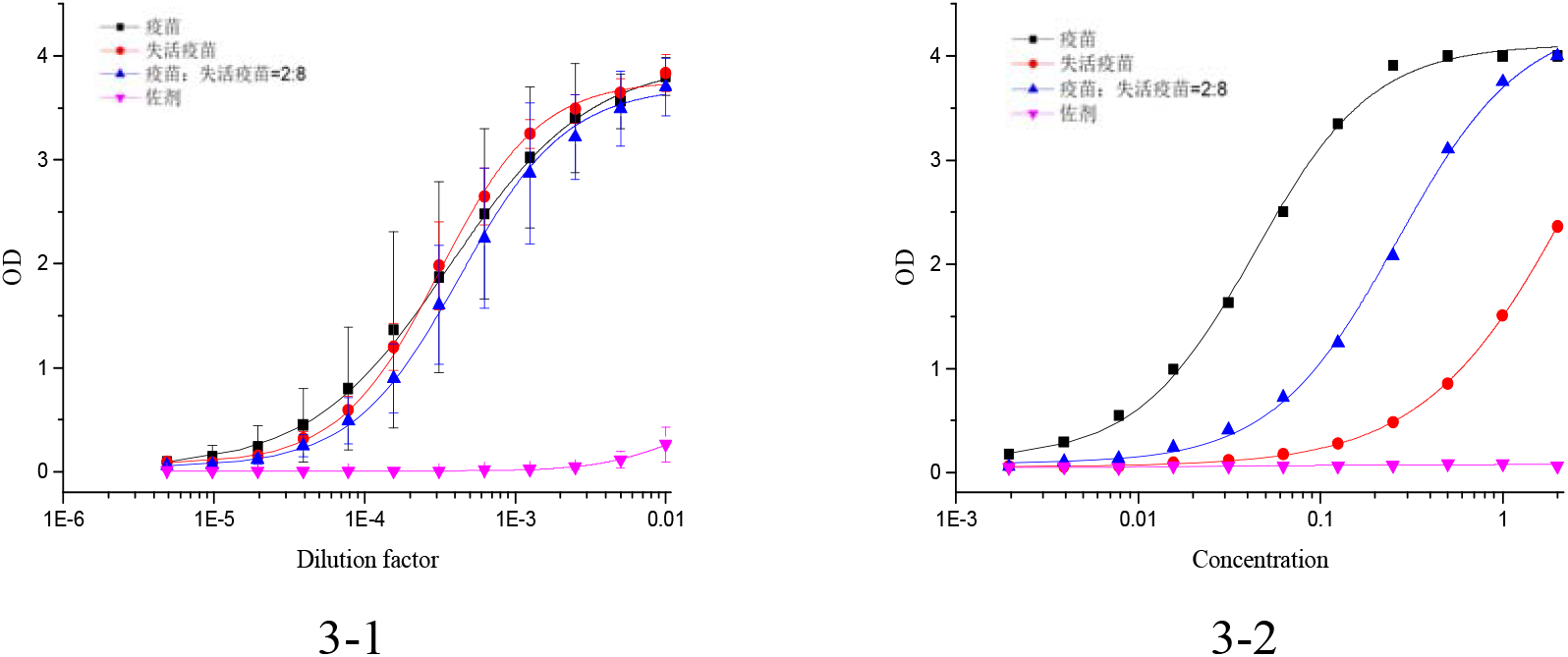
Heat treatment dramatically reduces the ELISA titers

### 3.5 Dissociate VLPs from adjuvant

To dissociate VLPs from aluminum hydroxyphosphate sulfate adjuvant, HPV VLP vaccines were incubated overnight at room temperature with continuous shaking in citrate-phosphate buffer (60mM sodium phosphate, 0.1M sodium citrate, 1M sodium chloride, 0.4% Tween 80, pH 6.7-6.8). The extracted VLPs were then analyzed by our HPV ELISA kit.

As shown in Figure 4, after the treatment of dissociation of adjuvant, most of the ELISA signals were persevered. Although the recovery rates for different HPV subtype VLPs varied from 70% to nearly 100%, the recovery rates for each subtype were very consistent between different tests, time points, and technicians.

**Figure 4.**
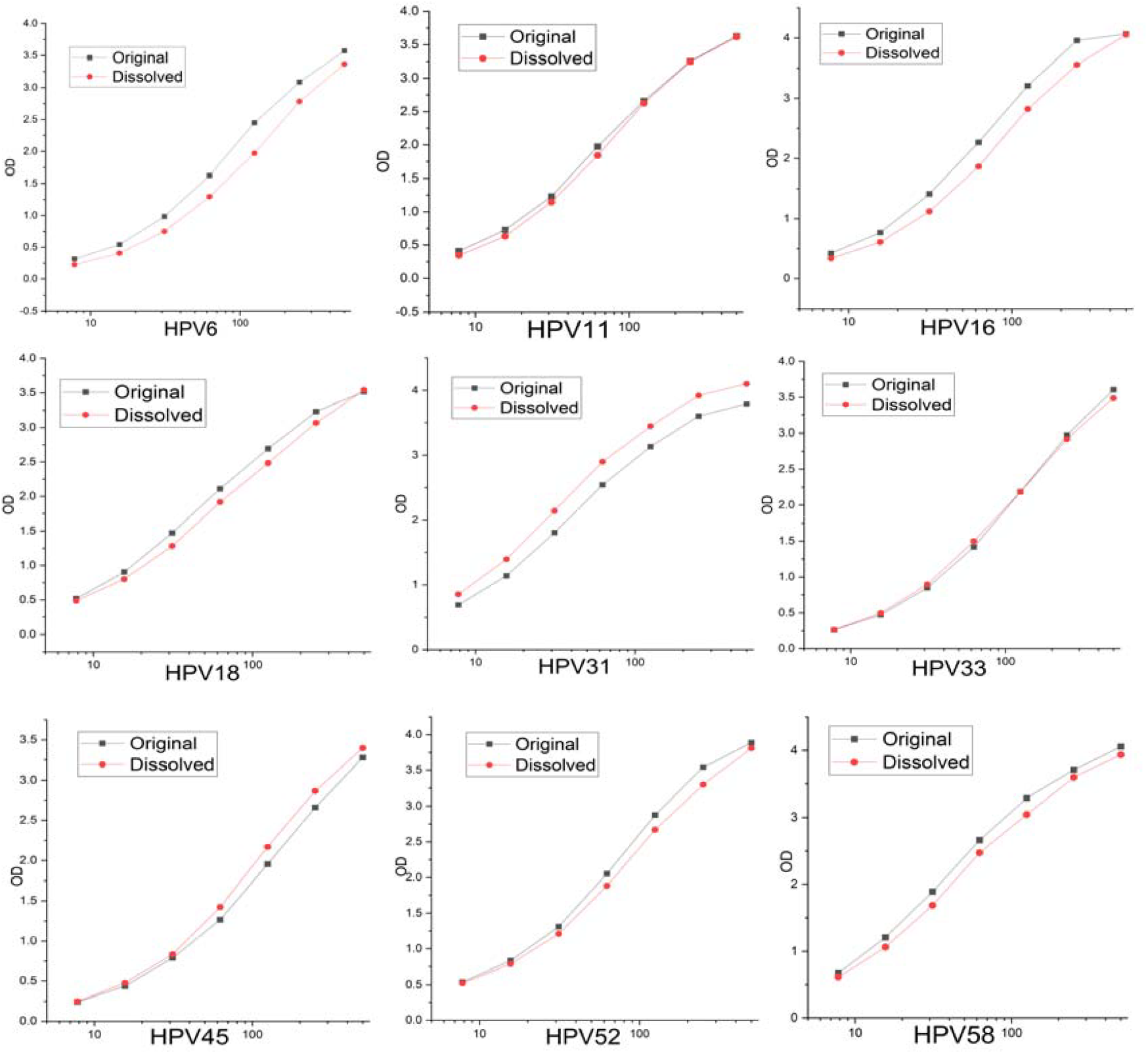
VLP detection of original and adjuvant-dissolved components from vaccine. Original as pre-mixed components and Dissolved as the VLP dissolved from adjuvants.

### 3.6 Broad reactivity towards HPV VLPs from different manufactueres

HPV VLPs can be produced using many different expression systems, such as yeast, bacteria and insect SF9 cells, and through different manufacture methods with or without disassembling/re-assembling. To access the suitability of our newly developed 9 HPV subtype-specific Sandwich ELISA kits, we tested them against different HPV VLPs from 6 different vaccine manufacturers in China. representing a diverse range of production systems and methods.

As shown in Fig 5, the 9 kits could measure the HPV VLPs from 6 different manufacturers, demonstrating the broad suitability. However, the result for HPV16 were highly consistent across 4 different manufacturers, while the data for HPV18 showed greater variability.

**Figure 5:**
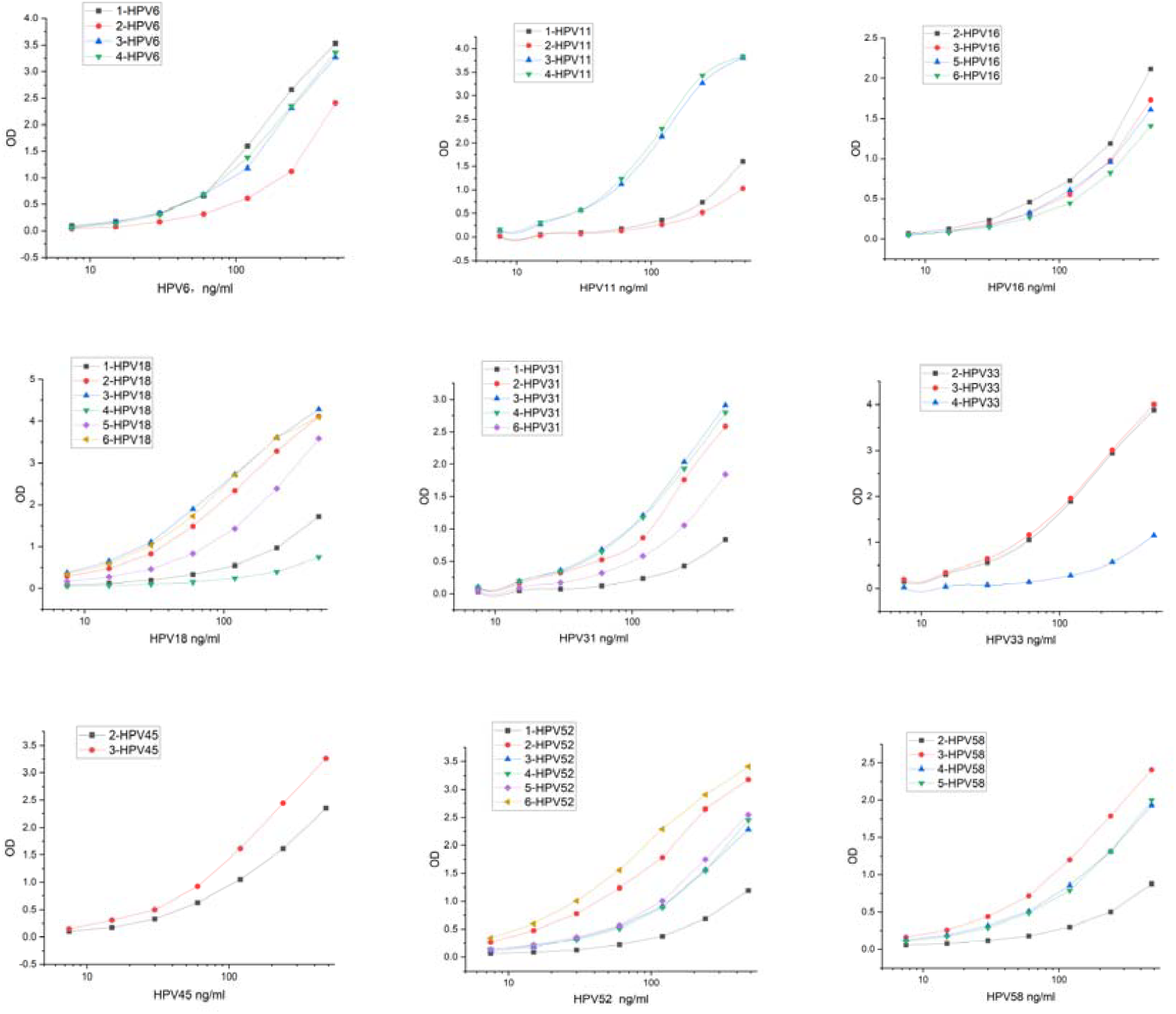
Broad-spectrum validation of subtype-specific HPV antibody pairs.

## 4. Discussion

CC represents a major global health burden for women, with approximately 530,000 new cases and 275,000 deaths annually. High-risk HPV infection, especially HPV 16 and 18, plays a pivotal role in cervical carcinogenesis. First-generation HPV vaccines targeted HPV 16/18, while newer vaccines provide broader protection against additional high-risk types. The recently developed 9-valent HPV vaccine had potential to prevent up to 90% of HPV-associated anogenital cancers. Clinical trials data showed efficacy protecting the CC through 70% - 100%. CC patients will tromentically drop after the immunization with HPV vaccine.

The efficacy of HPV vaccine is critical for manufacturers and recipients. In this study, we generated HPV subtype-specific monoclonal antibodies and developed quantitative detection kit to evaluate the quality of the 9 valent HPV vaccine. From the result above we can see our kit has perfect specificity to individual components of the VPLs, good lineage relationship for quantitative detection, good sensitivity to satisfy evaluation requirement(3.9-4000 ng/mL), and well compatibility with different vaccines from multiple manufacturers. It is also user-friendly and time-efficient.

The recently approved 9-valent HPV vaccine is being widely implemented globally as a critical tool for effective cervical cancer prevention. In addition to Merck, numerous manufacturers have developed or are developing HPV vaccines, which undergo rigorous efficacy and safety evaluation through clinical trials prior to market authorization. However, as biological products, inherent batch-to-batch variability is expected due to differences in raw materials, production systems, and cell line performance.

To enable accurate assessment of this variability and allow regulators to reliably evaluate diverse products, quantitative analytical methods to determine vaccine potency and immunogenicity in vitro are essential. And the accurate measurement of the key active VLP components in final formulations remains challenging.

VLPs represent the primary immunogenic component in current HPV vaccines. Assessing VLP integrity and conformational epitope exposure is central to evaluating vaccine potency in vitro. We demonstrated our ELISA can distinguish between active and heat-inactivated VLP samples, verifying selective detection of neutralizing epitopes that correlate with vaccine efficacy. Thus, the assays can provide both quantitative VLP content measurement and inform on VLP quality and immunogenicity through conformational epitope recognition.

Furthermore, HPV vaccine formulations contain various adjuvants, which differ across manufacturers. The ability to selectively detect VLPs in these complex heterogeneous mixtures is an unmet need. We have developed ELISAs that can accurately quantify individual VLPs in aluminum adjuvant-containing formulations following de-adjuvanting treatment. The assays overcome adjuvant interference, providing a critically needed standardized analytical toolkit for vaccine quality control and comparative potency assessment.

## Data Availability

All data produced in the present work are contained in the manuscript.

